# Risk factors for hospitalization, disease severity and mortality in children and adolescents with COVID-19: Results from a nationwide German registry

**DOI:** 10.1101/2021.06.07.21258488

**Authors:** J Armann, M Doenhardt, M Hufnagel, N Diffloth, F Reichert, W Haas, J Schilling, S Haller, J Hübner, A Simon, DT Schneider, J Brunner, A Trotter, M Roessler, J Schmitt, R Berner

## Abstract

**Objective:** To characterize the clinical features of children and adolescents hospitalized with SARS-CoV-2 infections and to explore predictors for disease severity.

**Design:** Nationwide prospective observational cohort study.

**Setting:** Data collected from 169 out of 351 children’s hospitals in Germany between March 18, 2020 and April 30, 2021 and comparison with the Statutory Notification System.

**Participants:** 1,501 children and adolescents up to 19 years of age with laboratory confirmed SARS-CoV-2 infections who were admitted to children’s hospitals and subsequently reported to the COVID-19 registry of the German Pediatric Infectious Disease Society (DGPI).

**Main outcome measures:** Admission to intensive care, in-hospital.

**Results:** As compared to the information in the statutory notification system, up to 30% of all children and adolescents hospitalized in Germany during the study period were reported to the DGPI registry. Median age was three years (IQR, 0-12), with 36% of reported cases being infants. Although roughly half of patients in the registry were not admitted to the hospital due to their SARS-CoV-2 infection, 72% showed infection-related symptoms during hospitalization. Preexisting comorbidities were present in 28%, most commonly respiratory disorders, followed by neurological, neuromuscular, and cardiovascular diseases. Median length of hospitalization was five days (IQR 3-10). Only 20% of patients received a SARS-CoV-2-related therapy. Infants were less likely to require therapy as compared to older children. Overall, 111 children and adolescents were admitted to intensive care units (ICU). In a fully adjusted model, patient age, trisomy 21, coinfections and primary immunodeficiencies (PID) were significantly associated with intensive care treatment. In a bivariate analysis, pulmonary hypertension, cyanotic heart disease, status post (s/p) cardiac surgery, fatty liver disease, epilepsy and neuromuscular impairment were statistically significant risk factors for ICU admission.

**Conclusion:** Overall, a small proportion of children and adolescents was hospitalized in Germany during the first year of the pandemic. The majority of patients within our registry was not admitted due to COVID-19 suggesting an overestimation of the disease burden even in hospitalized children. Nevertheless, a large proportion of children and adolescents with confirmed COVID-19 reported in Germany could be captured. This allowed for detailed assessment of overall disease severity and underlying risk factors in our cohort. The main risk factors for COVID-19 disease associated intensive care treatment were older patient age, trisomy 21, PIDs and coinfection at the time of hospitalization.

**Trial registration:** Registry of hospitalized pediatric patients with SARS-CoV-2 infection (COVID-19), DRKS00021506

## INTRODUCTION

Since December 2019, Severe Acute Respiratory Syndrome Coronavirus 2 (SARS-CoV-2) [1] has rapidly spread and emerged as a global pandemic. The role of children and adolescents in the pandemic has been debated within the scientific community and beyond. Although the pediatric population — in contrast to adults [2] — usually has a mild disease course and low hospitalization rates [3, 4], there are still children who experience severe disease [5] and who therefore would benefit from non-pharmaceutical protective measures and vaccination. For this reason, it is critical to identify populations at risk within the pediatric population in order to implement preventive measures targeted to reduce COVID-19-associated morbidity and mortality for vulnerable children and adolescents, to focus interventions thereby preventing potential harm to the pediatric population as a whole when imposing restrictive mitigation measures.

Since March 18, 2020, the German Society for Pediatric Infectious Diseases (DGPI), supported by several additional German professional pediatric societies, has collected nationwide data on children and adolescents with laboratory confirmed SARS-CoV-2 infections who were admitted to pediatric departments and hospitals in Germany. With this clinical registry of 1,501 pediatric patients, we are able to provide reliable, detailed information on clinical characteristics, disease course and outcome predictors among children and adolescents.

## METHODS

We implemented a national, prospective registry for children and adolescents hospitalized with a SARS-CoV-2 infection. The registry was approved by the Ethics Committee of the Technische Universität (TU) Dresden (BO-EK-110032020) and was assigned clinical trial number DRKS00021506.

### Settings and case definitions

All German pediatric hospitals, along with members of the German scientific pediatric and pediatric infectious diseases societies, (German Society of Pediatric Infectious Diseases (DGPI), German Society of Pediatrics and Adolescents Medicine (DGKJ), Association of Senior Pediatricians and Pediatric Surgeons in Germany (VLKKD)), were invited to participate. Patients who had laboratory-confirmed SARS-CoV-2 infections and who were hospitalized between March 18, 2020 and April 30, 2021 were eligible. 169 Hospitals in Germany reported cases. Reporting of children admitted to hospitals from Austria was possible, 2 hospitals did.

Confirmed SARS-CoV-2 infection was defined either by a positive real-time reverse transcriptase polymerase chain reaction (RT-PCR) test or else by a rapid antigen test (RAT) for SARS-CoV-2 which were deemed positive by the reporting physician.

For each patient, an electronic case report form was completed with an access point via the DGPI website (https://dgpi.de/covid-19-survey-der-dgpi/) that linked to REDCap electronic data capture tools hosted at TU Dresden. [6, 7] REDCap (Research Electronic Data Capture) is a secure, web-based software platform designed to support data capture for research studies, providing 1) an intuitive interface for validated data capture; 2) audit trails for tracking data manipulation and export procedures; 3) automated export procedures for seamless data downloads to common statistical packages; and 4) procedures for data integration and interoperability with external sources.

Data collected by a predefined list (see above) included demographic characteristics, exposure, comorbidities, initial symptoms and clinical signs, treatments, disease course during hospitalization, and outcome at discharge from the hospital.

We additionally analyzed data from the statutory notification system of laboratory-confirmed SARS-CoV-2 infections in Germany between March 18, 2020 and April 30, 2021 for patients <18 years of age. In Germany, laboratory confirmation requires detection of SARS-CoV-2 nucleic acid by PCR or culture isolation of the pathogen (according to the national case definition [8]. Physicians and laboratories have to notify cases to the local public health authorities (PHA) who forward the details via the respective state PHA to the Robert Koch Institute (RKI, national public health institute) in Berlin. Reporting includes information on symptoms, hospitalization status and pre-existing conditions. Incidences were calculated as 7-day incidence or cumulative incidence over the entire observation period (18 March 2020 – 30 April 2021) using population estimates for 31 December 2019 [9].

Comparison of the two data sets allowed us to estimate the coverage extent of our registry.

### Comorbidities

The following groups of comorbidities were considered to be possible risk factors for severe course of SARS-CoV-2 infection: respiratory, cardiovascular, gastrointestinal, liver, renal, neurological/neuromuscular, psychiatric, hematological, oncological, autoimmune, or syndromic diseases, as well as primary immunodeficiencies (PIDs), s/p transplant (solid organ or bone marrow), history of prematurity, tracheostomy and/or home oxygen prior to SARS-CoV-2 infection.

In addition, diagnoses with an occurrence in at least n=7 patients were analyzed in a bivariate model (i.e., asthma/wheezing, cyanotic and acyanotic heart disease, hypertension, fatty liver disease, epilepsy, neuromuscular impairment, diabetes, acute leukemia, hemolytic anemia and s/p cardiac surgery).

### Outcome Measure

Among included cases, the main outcome categories tracked were: symptomatic infection, case fatality, sequelae at discharge and severe disease, as defined by the requirement for intensive care unit (ICU) admission due to COVID19.

### Statistical Analysis

Analyses were performed using IBM SPSS v.25.0, R v.3.6.3 and Microsoft Excel v.2010. We present descriptive statistics as medians with first and third quartiles for continuous variables and as absolute frequencies with percentages for categorical variables. Using robust Poisson regression [10], we estimated relative risk for severe disease evolution in terms of ICU admission. In addition to bivariate models for ICU admission and specific risk factors, we estimated a fully adjusted model which included all considered risk factors simultaneously. We quantified the precision of relative risk estimates by 95% confidence intervals (CI) and applied a significance level of 0.05 two-sided.

## RESULTS

### Demographics and clinical characteristics – statutory notification system

From March 18, 2020 to April 30, 2021, 417,527 children and adolescents <18 years with laboratory-confirmed SARS-CoV-2 infection were reported to the RKI. The median age of all reported cases was 10 years (IQR 6-14).

Among cases with information on hospitalization status, 1.5% (4,921/320,360) were hospitalized (Table 2). The median age of hospitalized cases was seven years (IQR 1-14). The weekly number of children and adolescents hospitalized peaked End of March 2020 (week 13, n = 57), in December 2020 (week 51, n = 210) and April 2020 (week 15, n =219). This mirrored incidence rates in the general population and of the three COVID-19 waves in Germany (Fig.1).

**Figure 1.**
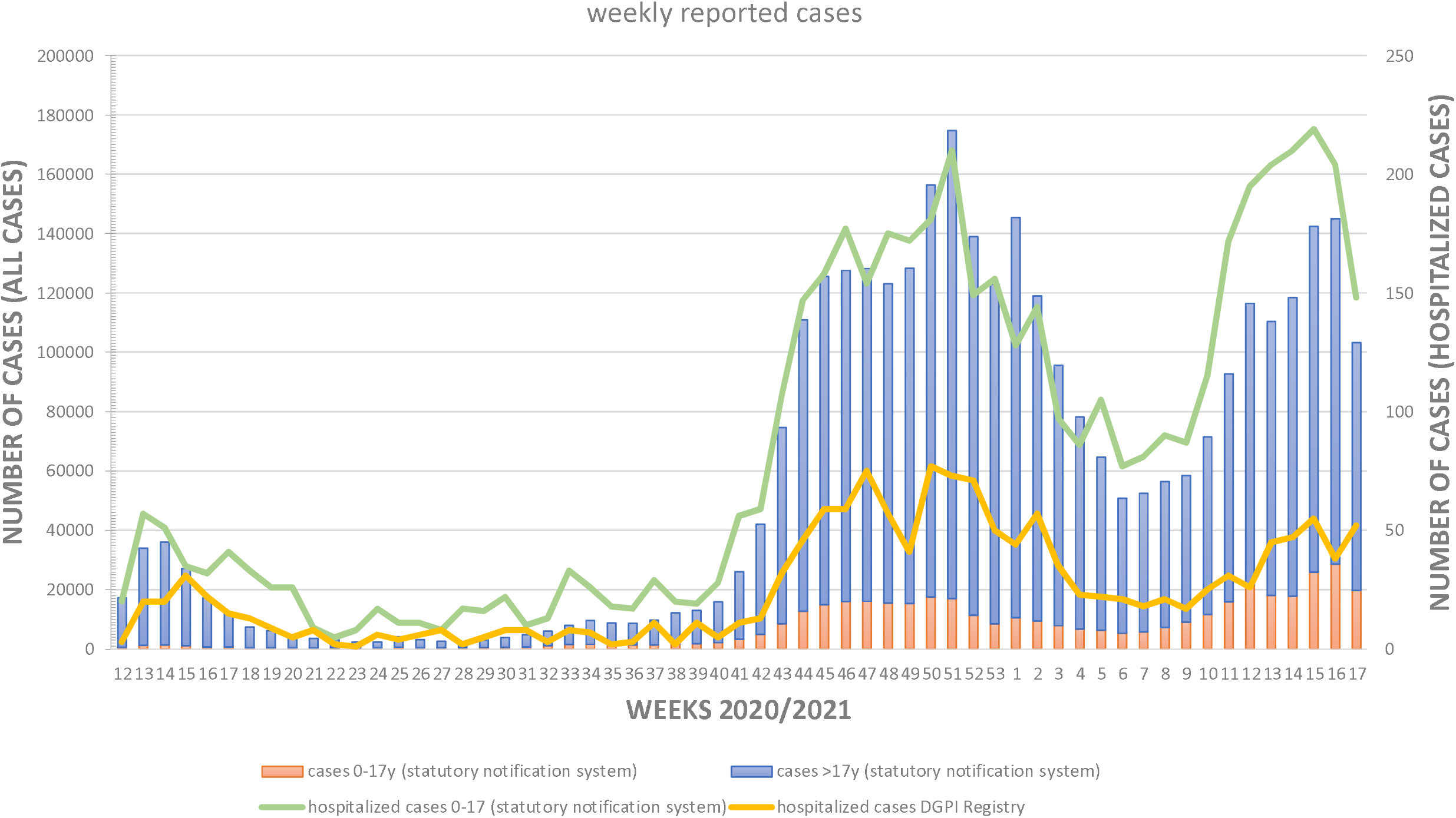
Weekly reported SARS-CoV-2 cases by age group and hospital admissions. Weeks in the statutory reporting system refer to reporting weeks, weeks in the DGPI Registry refer to admission date

**Table 1.**
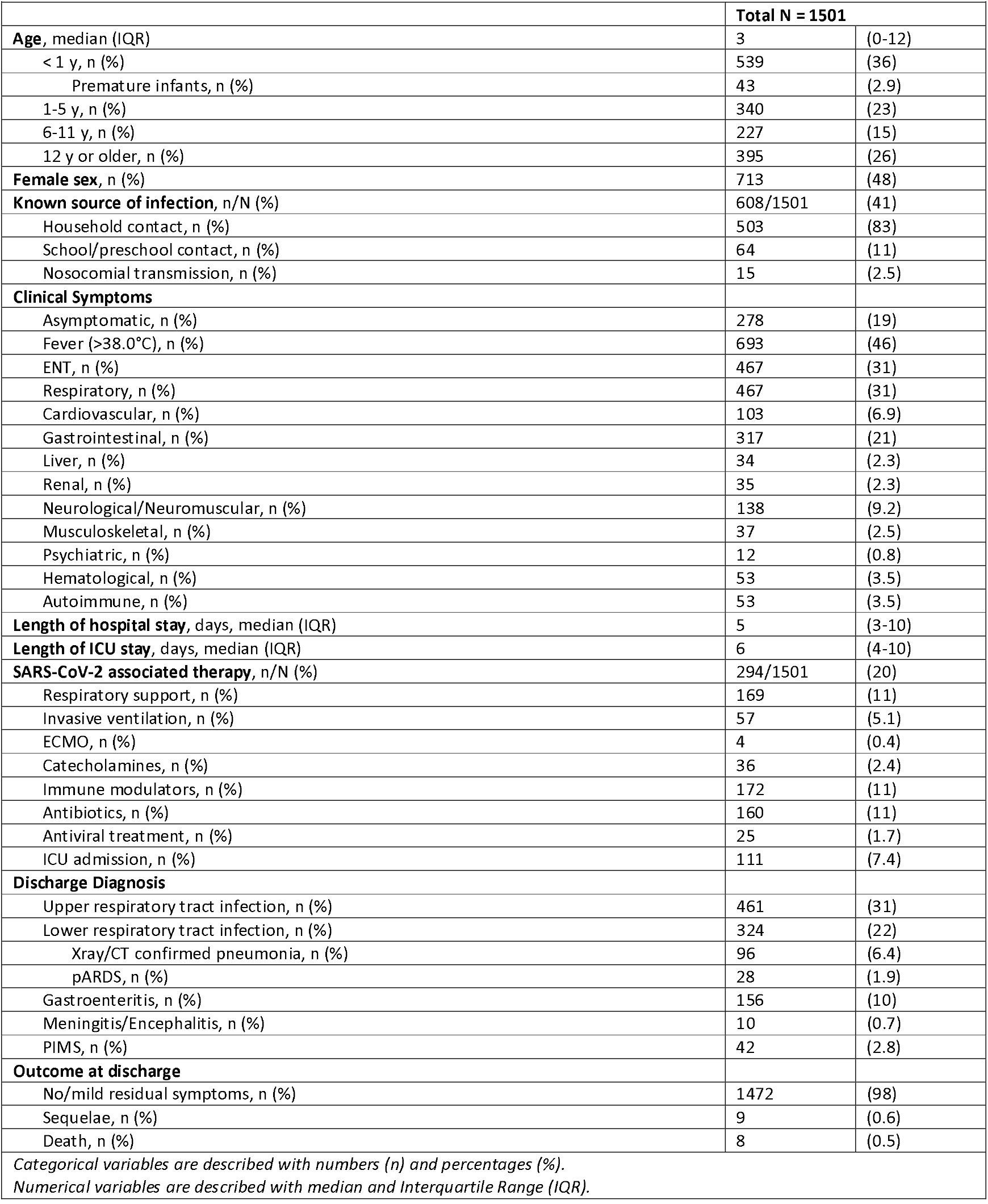
**General characteristics of hospitalized children and adolescents with a laboratory-confirmed SARS-CoV-2 infection**

|  |  |  |
| --- | --- | --- |
|  | **Total N = 1501** |  |
| **Age, median (IQR)** | 3 | (0-12) |
| < 1 y, n (%) | 539 | (36) |
| Premature infants, n (%) | 43 | (2.9) |
| 1-5 y, n (%) | 340 | (23) |
| 6-11 y, n (%) | 227 | (15) |
| 12 y or older, n (%) | 395 | (26) |
| **Female sex, n (%)** | 713 | (48) |
| **Known source of infection, n/N (%)** | 608/1501 | (41) |
| Household contact, n (%) | 503 | (83) |
| School/preschool contact, n (%) | 64 | (11) |
| Nosocomial transmission, n (%) | 15 | (2.5) |
| **Clinical Symptoms** |  |  |
| Asymptomatic, n (%) | 278 | (19) |
| Fever (>38.0°C), n (%) | 693 | (46) |
| ENT, n (%) | 467 | (31) |
| Respiratory, n (%) | 467 | (31) |
| Cardiovascular, n (%) | 103 | (6.9) |
| Gastrointestinal, n (%) | 317 | (21) |
| Liver, n (%) | 34 | (2.3) |
| Renal, n (%) | 35 | (2.3) |
| Neurological/Neuromuscular, n (%) | 138 | (9.2) |
| Musculoskeletal, n (%) | 37 | (2.5) |
| Psychiatric, n (%) | 12 | (0.8) |
| Hematological, n (%) | 53 | (3.5) |
| Autoimmune, n (%) | 53 | (3.5) |
| **Length of hospital stay, days, median (IQR)** | 5 | (3-10) |
| **Length of ICU stay, days, median (IQR)** | 6 | (4-10) |
| **SARS-CoV-2 associated therapy, n/N (%)** | 294/1501 | (20) |
| Respiratory support, n (%) | 169 | (11) |
| Invasive ventilation, n (%) | 57 | (5.1) |
| ECMO, n (%) | 4 | (0.4) |
| Catecholamines, n (%) | 36 | (2.4) |
| Immune modulators, n (%) | 172 | (11) |
| Antibiotics, n (%) | 160 | (11) |
| Antiviral treatment, n (%) | 25 | (1.7) |
| ICU admission, n (%) | 111 | (7.4) |
| **Discharge Diagnosis** |  |  |
| Upper respiratory tract infection, n (%) | 461 | (31) |
| Lower respiratory tract infection, n (%) | 324 | (22) |
| Xray/CT confirmed pneumonia, n (%) | 96 | (6.4) |
| pARDS, n (%) | 28 | (1.9) |
| Gastroenteritis, n (%) | 156 | (10) |
| Meningitis/Encephalitis, n (%) | 10 | (0.7) |
| PIMS, n (%) | 42 | (2.8) |
| **Outcome at discharge** |  |  |
| No/mild residual symptoms, n (%) | 1472 | (98) |
| Sequelae, n (%) | 9 | (0.6) |
| Death, n (%) | 8 | (0.5) |
| *Categorical variables are described with numbers (n) and percentages (%). Numerical variables are described with median and Interquartile Range (IQR).* |  |  |

**Table 2.** **Statutory notification system (RKI) vs. DGPI Registry: SARS-CoV-2 associated symptoms, therapy, hospitalization and ICU admission**

|  |  |  |  |  |  |  |
| --- | --- | --- | --- | --- | --- | --- |
| **Statutory notification system (RKI)** |  |  |  |  |  |  |
|  | **< 1 y** | **1-17 y** | **p** | **1-5 y** | **6-11 y** | **12-17 y** |
| SARS-CoV-2 associated symptoms, n/N (%) * | 6,772/<br>8,863<br>(76) | 185,061/<br>269,361<br>(69) | <0.01 | 37,146/<br>58,862<br>(63) | 53,811/<br>86,815<br>(62) | 94,104/<br>123,684<br>(76) |
| Hospitalization n/N (%) ** | 1,157/<br>10,436<br>(11) | 3,764/<br>309,924<br>(1.2) | <0.01 | 1,050/<br>70,258<br>(1.5) | 941/<br>102,457<br>(0.9) | 1773/<br>137,209<br>(1.3) |
| ICU admission n/N (%) *** | 21/<br>1,022<br>(2.1) | 46/<br>2,273<br>(2.0) | NS | 9/<br>716<br>(1.3) | 11/<br>475<br>(2.3) | 26/<br>1,082<br>(2.4) |
| **DGPI Registry** |  |  |  |  |  |  |
|  | **< 1 y** | **$\geq 1$ y** | **p** | **1-5 y** | **6-11 y** | **$\geq 12$ y** |
| SARS-CoV-2 associated symptoms, n/N (%) | 428/<br>539<br>(79) | 628/<br>962<br>(65) | <0.01 | 235/<br>340<br>(69) | 131/<br>227<br>(58) | 262/<br>395<br>(66) |
| SARS-CoV-2 associated therapy, n/N (%) | 76/<br>539<br>(14) | 218/<br>962<br>(23) | <0.01 | 68/<br>340<br>(18) | 57/<br>227<br>(19) | 93/<br>395<br>(18) |
| ICU admission n/N (%) | 12/<br>539<br>(2.2) | 99/<br>962<br>(10) | <0.01 | 19/<br>340<br>(5.6) | 36/<br>227<br>(16) | 44/<br>395<br>(11) |
| * All cases (regardless of hospitalization status) for which information regarding symptoms is reported (N= 278,224) |  |  |  |  |  |  |
| ** All cases for which hospitalization status is available (N=320,360) |  |  |  |  |  |  |
| *** All cases for which ICU status is available (N=3,295) |  |  |  |  |  |  |

For 278,224 (66.6%) and 3,295 (0.8% of all and 67% of hospitalized) cases, reporting included information on presence on symptoms and ICU status, respectively. Incidence of infections and symptomatic infections by age group was highest among 12- to 17-year-old children. However, hospitalizations per 100,000 population were highest among infants (Fig.2).

**Figure 2.**
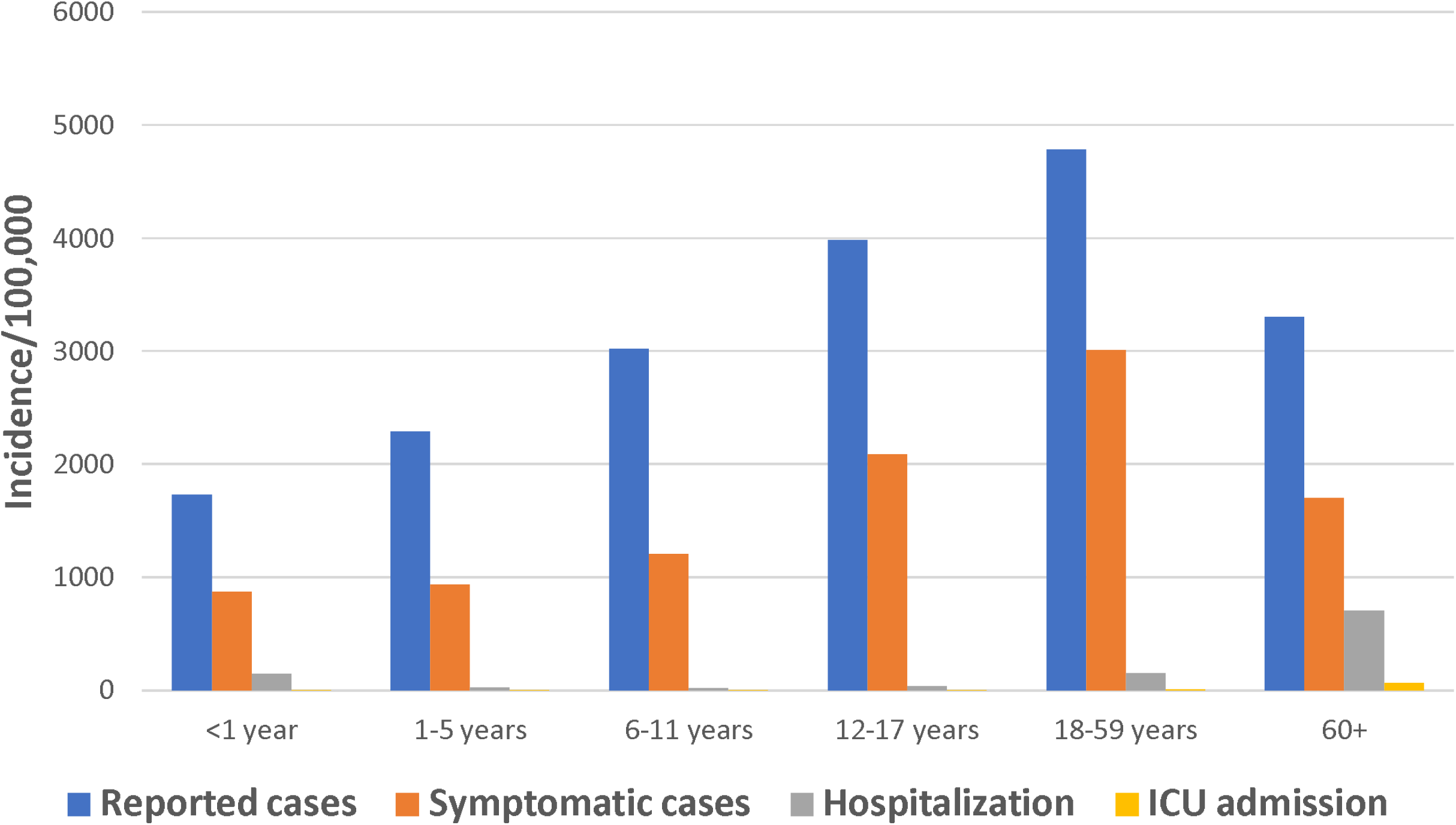
Cumulative incidence of infections, symptomatic infections, hospitalizations, and ICU admissions by age group (statutory notification system), 18 March 2020 – 30 April 2021.

Among cases with corresponding information comorbidities were reported in 3.7% (8,786/239,093) of all and 11.1% (372/3,354) of hospitalized cases. The most frequently reported comorbidity was respiratory disease, followed by cardiovascular and renal disease. The proportion of hospitalizations was higher among children with comorbidities, with the exception of patients with pre-existing respiratory disease (Supplemental Table 1).

### Demographics and clinical characteristics – DGPI Registry

During the same period, 1,501 hospitalized children and adolescents with laboratory confirmed SARS-CoV-2 infection (n= 1,492 RT-PCR; n= 9 RAT) were reported via the DGPI registry. Of these, 1,473 were hospitalized in Germany with 169/351 (48%) of all children’s hospitals or pediatric departments reporting cases from all 16 federal states. This corresponds to 30% of all children with known hospitalization status in the statutory notification system in this age group; An additional 28 patients were reported from hospitals in Austria. In the DGPI registry, median age was three years (IQR, 0-12), with 36% of reported cases being infants < 1 year of age. Of these, 43 were born prematurely, with a median gestational age of 35 weeks (range 23-36). There was no gender predominance (48% female).

In 608/1,501 (41%) cases, a known source of infection was reported, most commonly (83%) household contacts, followed by school and preschool contacts (11%). Nosocomial transmission was documented in 2.5% of cases.

Of the 1,501 cases, 806 (54%) were admitted to the hospital not for their SARS-CoV-2 infection, but obviously for other reasons. In 72% of patients (n=1,084), SARS-CoV-2-associated symptoms during hospitalization were reported. By contrast, 19% (n=278) were asymptomatic and 9% (n=139) had symptoms that could not with certainty be assigned to their SARS-CoV-2 infection. Fever was the most commonly reported symptom (46%), followed by ear, nose and throat (ENT) (31%), respiratory (31%) and gastrointestinal symptoms (21%). Upper respiratory tract infection was the most common diagnosis at discharge (31%), followed by lower respiratory tract infection (22% overall, including 8.3% (n=125) with pneumonia and 1.9% (n=28) with pediatric acute respiratory distress syndrome (pARDS)). Median age of patients with pARDS was nine years (IQR 6-15). In 5% (n= 42) of cases with a PCR-confirmed infection, diagnosis of Pediatric Inflammatory Multisystem Syndrome (PIMS), also referred to as Multisystem Inflammatory Syndrome in Children (MIS-C), was made.

Preexisting comorbidities were present in 419 cases (28%). Most commonly these were respiratory disorders (n=103), followed by neurological and neuromuscular disorders (n=101) and cardiovascular disorders (n=74) (Table 3).

**Table 3.**
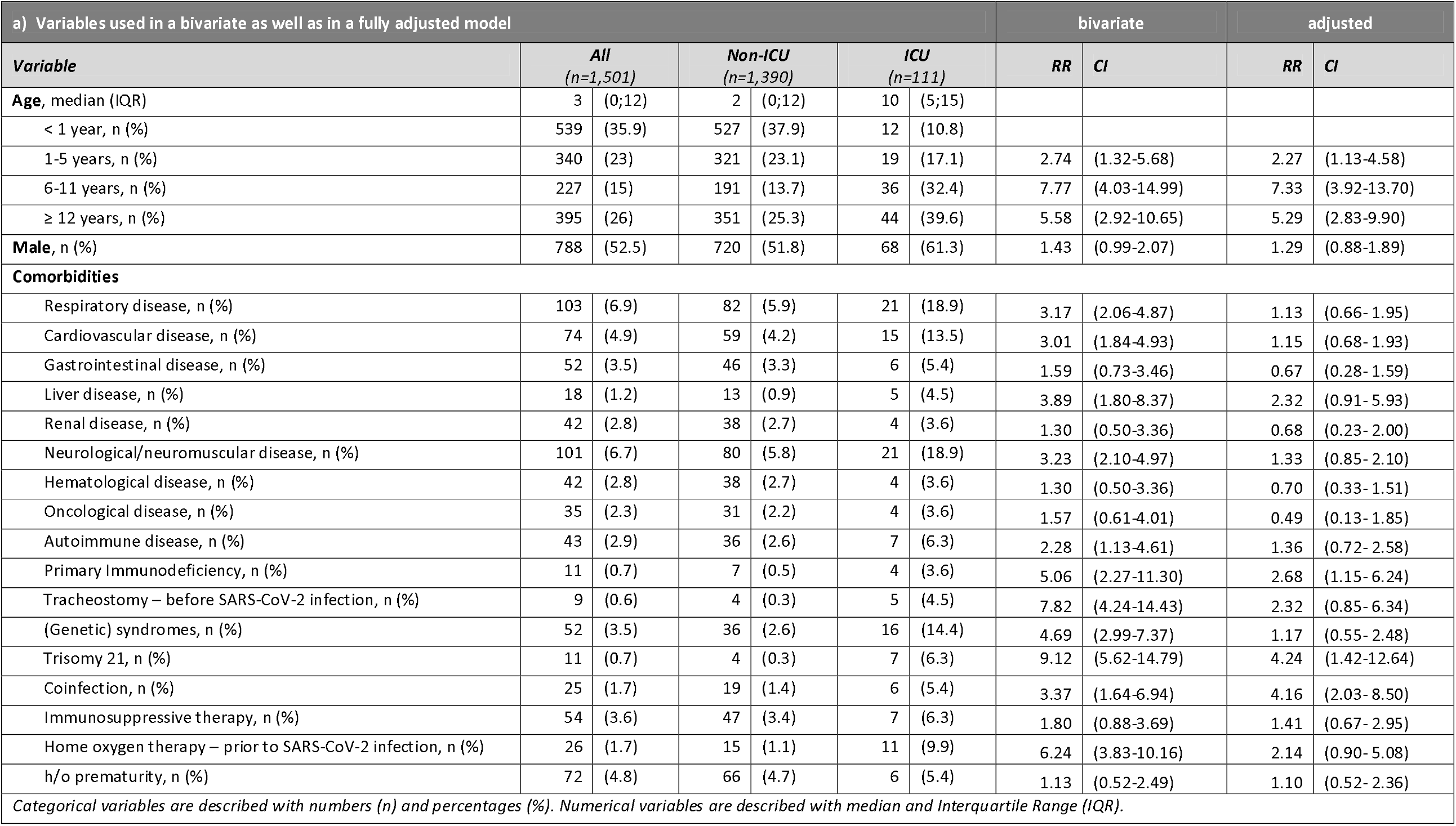
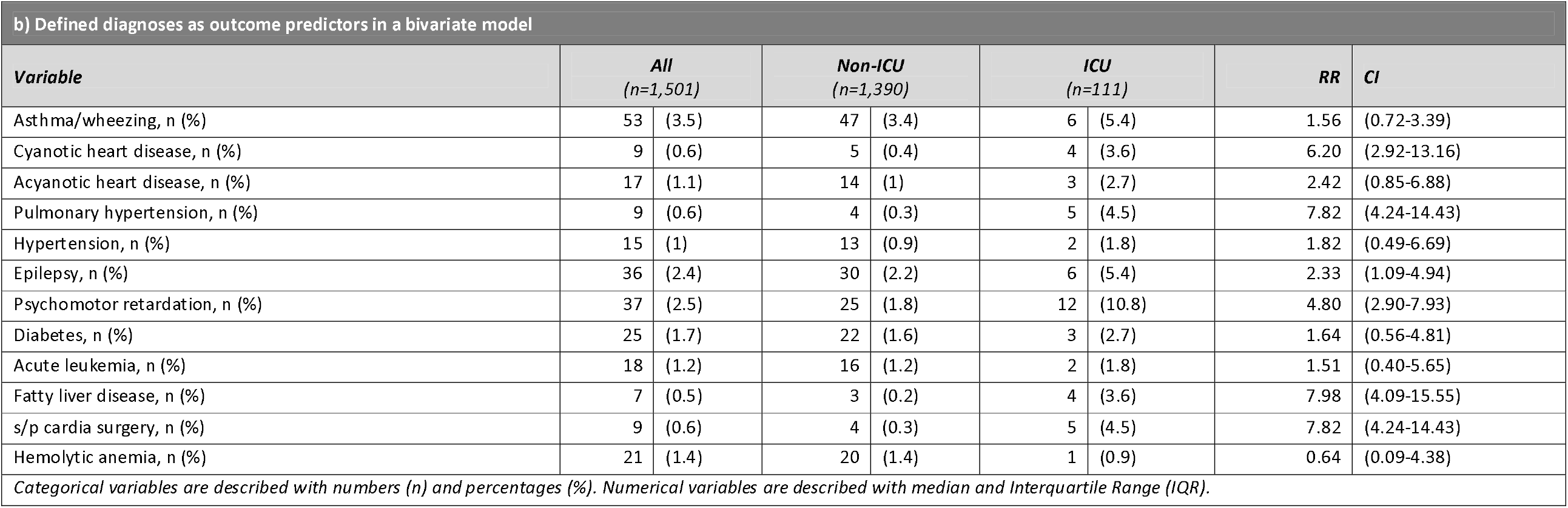
Outcome and disease severity predictors.

| a) Variables used in a bivariate as well as in a fully adjusted model |  |  |  |  |  |  | bivariate |  | adjusted |  |
| --- | --- | --- | --- | --- | --- | --- | --- | --- | --- | --- |
| Variable | All<br>(n=1,501) |  | Non-ICU<br>(n=1,390) |  | ICU<br>(n=111) |  | RR | CI | RR | CI |
| Age, median (IQR) | 3 | (0;12) | 2 | (0;12) | 10 | (5;15) |  |  |  |  |
| < 1 year, n (%) | 539 | (35.9) | 527 | (37.9) | 12 | (10.8) |  |  |  |  |
| 1-5 years, n (%) | 340 | (23) | 321 | (23.1) | 19 | (17.1) | 2.74 | (1.32-5.68) | 2.27 | (1.13-4.58) |
| 6-11 years, n (%) | 227 | (15) | 191 | (13.7) | 36 | (32.4) | 7.77 | (4.03-14.99) | 7.33 | (3.92-13.70) |
| ≥ 12 years, n (%) | 395 | (26) | 351 | (25.3) | 44 | (39.6) | 5.58 | (2.92-10.65) | 5.29 | (2.83-9.90) |
| Male, n (%) | 788 | (52.5) | 720 | (51.8) | 68 | (61.3) | 1.43 | (0.99-2.07) | 1.29 | (0.88-1.89) |
| **Comorbidities** |  |  |  |  |  |  |  |  |  |  |
| Respiratory disease, n (%) | 103 | (6.9) | 82 | (5.9) | 21 | (18.9) | 3.17 | (2.06-4.87) | 1.13 | (0.66- 1.95) |
| Cardiovascular disease, n (%) | 74 | (4.9) | 59 | (4.2) | 15 | (13.5) | 3.01 | (1.84-4.93) | 1.15 | (0.68- 1.93) |
| Gastrointestinal disease, n (%) | 52 | (3.5) | 46 | (3.3) | 6 | (5.4) | 1.59 | (0.73-3.46) | 0.67 | (0.28- 1.59) |
| Liver disease, n (%) | 18 | (1.2) | 13 | (0.9) | 5 | (4.5) | 3.89 | (1.80-8.37) | 2.32 | (0.91- 5.93) |
| Renal disease, n (%) | 42 | (2.8) | 38 | (2.7) | 4 | (3.6) | 1.30 | (0.50-3.36) | 0.68 | (0.23- 2.00) |
| Neurological/neuromuscular disease, n (%) | 101 | (6.7) | 80 | (5.8) | 21 | (18.9) | 3.23 | (2.10-4.97) | 1.33 | (0.85- 2.10) |
| Hematological disease, n (%) | 42 | (2.8) | 38 | (2.7) | 4 | (3.6) | 1.30 | (0.50-3.36) | 0.70 | (0.33- 1.51) |
| Oncological disease, n (%) | 35 | (2.3) | 31 | (2.2) | 4 | (3.6) | 1.57 | (0.61-4.01) | 0.49 | (0.13- 1.85) |
| Autoimmune disease, n (%) | 43 | (2.9) | 36 | (2.6) | 7 | (6.3) | 2.28 | (1.13-4.61) | 1.36 | (0.72- 2.58) |
| Primary Immunodeficiency, n (%) | 11 | (0.7) | 7 | (0.5) | 4 | (3.6) | 5.06 | (2.27-11.30) | 2.68 | (1.15- 6.24) |
| Tracheostomy – before SARS-CoV-2 infection, n (%) | 9 | (0.6) | 4 | (0.3) | 5 | (4.5) | 7.82 | (4.24-14.43) | 2.32 | (0.85- 6.34) |
| (Genetic) syndromes, n (%) | 52 | (3.5) | 36 | (2.6) | 16 | (14.4) | 4.69 | (2.99-7.37) | 1.17 | (0.55- 2.48) |
| Trisomy 21, n (%) | 11 | (0.7) | 4 | (0.3) | 7 | (6.3) | 9.12 | (5.62-14.79) | 4.24 | (1.42-12.64) |
| Coinfection, n (%) | 25 | (1.7) | 19 | (1.4) | 6 | (5.4) | 3.37 | (1.64-6.94) | 4.16 | (2.03- 8.50) |
| Immunosuppressive therapy, n (%) | 54 | (3.6) | 47 | (3.4) | 7 | (6.3) | 1.80 | (0.88-3.69) | 1.41 | (0.67- 2.95) |
| Home oxygen therapy – prior to SARS-CoV-2 infection, n (%) | 26 | (1.7) | 15 | (1.1) | 11 | (9.9) | 6.24 | (3.83-10.16) | 2.14 | (0.90- 5.08) |
| h/o prematurity, n (%) | 72 | (4.8) | 66 | (4.7) | 6 | (5.4) | 1.13 | (0.52-2.49) | 1.10 | (0.52- 2.36) |
| Categorical variables are described with numbers (n) and percentages (%). Numerical variables are described with median and Interquartile Range (IQR). |  |  |  |  |  |  |  |  |  |  |

**b) Defined diagnoses as outcome predictors in a bivariate model**
| **Variable** | **All**<br>**(n=1,501)** |  | **Non-ICU**<br>**(n=1,390)** |  | **ICU**<br>**(n=111)** |  | **RR** | **CI** |
| --- | --- | --- | --- | --- | --- | --- | --- | --- |
| Asthma/wheezing, n (%) | 53 | (3.5) | 47 | (3.4) | 6 | (5.4) | 1.56 | (0.72-3.39) |
| Cyanotic heart disease, n (%) | 9 | (0.6) | 5 | (0.4) | 4 | (3.6) | 6.20 | (2.92-13.16) |
| Acyanotic heart disease, n (%) | 17 | (1.1) | 14 | (1) | 3 | (2.7) | 2.42 | (0.85-6.88) |
| Pulmonary hypertension, n (%) | 9 | (0.6) | 4 | (0.3) | 5 | (4.5) | 7.82 | (4.24-14.43) |
| Hypertension, n (%) | 15 | (1) | 13 | (0.9) | 2 | (1.8) | 1.82 | (0.49-6.69) |
| Epilepsy, n (%) | 36 | (2.4) | 30 | (2.2) | 6 | (5.4) | 2.33 | (1.09-4.94) |
| Psychomotor retardation, n (%) | 37 | (2.5) | 25 | (1.8) | 12 | (10.8) | 4.80 | (2.90-7.93) |
| Diabetes, n (%) | 25 | (1.7) | 22 | (1.6) | 3 | (2.7) | 1.64 | (0.56-4.81) |
| Acute leukemia, n (%) | 18 | (1.2) | 16 | (1.2) | 2 | (1.8) | 1.51 | (0.40-5.65) |
| Fatty liver disease, n (%) | 7 | (0.5) | 3 | (0.2) | 4 | (3.6) | 7.98 | (4.09-15.55) |
| s/p cardia surgery, n (%) | 9 | (0.6) | 4 | (0.3) | 5 | (4.5) | 7.82 | (4.24-14.43) |
| Hemolytic anemia, n (%) | 21 | (1.4) | 20 | (1.4) | 1 | (0.9) | 0.64 | (0.09-4.38) |

*Categorical variables are described with numbers (n) and percentages (%). Numerical variables are described with median and Interquartile Range (IQR).*

### Treatment

Median length of hospitalization was five days (IQR 3-10). Only 20% (n=294) of patients received a SARS-CoV-2-related (defined by the physician who reported the case) therapy during their hospitalizations, most commonly immunomodulatory medication (11%), followed by respiratory support (11%) and antibiotics (11%). 111 patients (7.4%) were admitted to the intensive care unit (ICU) due to COVID-19. 57 (5.1%) were ventilated and 4 (0.4%) required Extracorporal Membrane Oxygenation (ECMO). Infants were less likely to receive a SARS-CoV-2-related therapy as compared to children >/= 1 year of age (14% vs. 23%; p 0.0001). Premature infants were just as likely to require therapy as infants overall (14% for both).

### Outcome and disease severity predictors

For 1,472 patients (98%), outcome was favorable. Upon discharge, they were either symptom-free or else had only mild, residual symptoms. Twelve patients (1%) were transferred to another hospital, and eight (0.7%) left the hospital with residual symptoms deemed to be long-term or permanent at the time of discharge. In total, eight (0.7%) patients died. Four of these deaths were COVID-19-associated, three previously were in palliative care due to a pre-existing disease, and one death could not be classified. Seven of the eight deceased patients had significant comorbidities (autoimmune, cardiovascular, neurological, gastrointestinal, primary immunodeficiencies (PIDs) and/or syndromes). Their age range was 6 months to 14 years; five were female.

Robust Poisson regression was used to model relationships between risk factors and ICU admission as a marker for disease severity (Table 3/Figure 3). Since no ICU patient was reported to have psychiatric disorders or to have received transplantation, these potential risk factors were excluded from the regression analyses. In a bivariate model, the following were significantly associated with a severe course of disease requiring ICU admission: age >1 year, pre-existing autoimmunological, cardiovascular, pulmonary, liver or neurological/neuromuscular disease, PID and syndromes, trisomy 21 as well as coinfection, pre-existing tracheostomy or home oxygen therapy. In the fully adjusted model, patient age, trisomy 21, coinfections and PIDs remained significantly associated with disease severity. Compared to infants (as reference), the estimated relative risk for ICU admission was 2.3 (95%-CI=1.1-4.6) times as high for children aged 1-5 years, 7.3 (95%-CI=3.9-13.7) for those aged 6-11, and 5.3 (95%-CI=2.8-9.9) for those over 12 years. Trisomy 21 was related to a 4.2-fold (95%-CI=1.4-12.6) higher risk, PID to a 2.7-fold (95%-CI=1.2-6.2) and coinfection to a 4.2 (2.0-8.5) higher risk as compared to children without these conditions. Bivariate analysis of defined diagnosis identified pulmonary hypertension, cyanotic heart disease, s/p cardiac surgery, fatty liver disease, epilepsy and neuromuscular impairment as statistically significant risk factors for ICU admission. However, asthma/wheezing, hypertension, acyanotic heart disease, diabetes, acute leukemia and hemolytic anemia did not appear to be risk factors.

**Figure 3.**
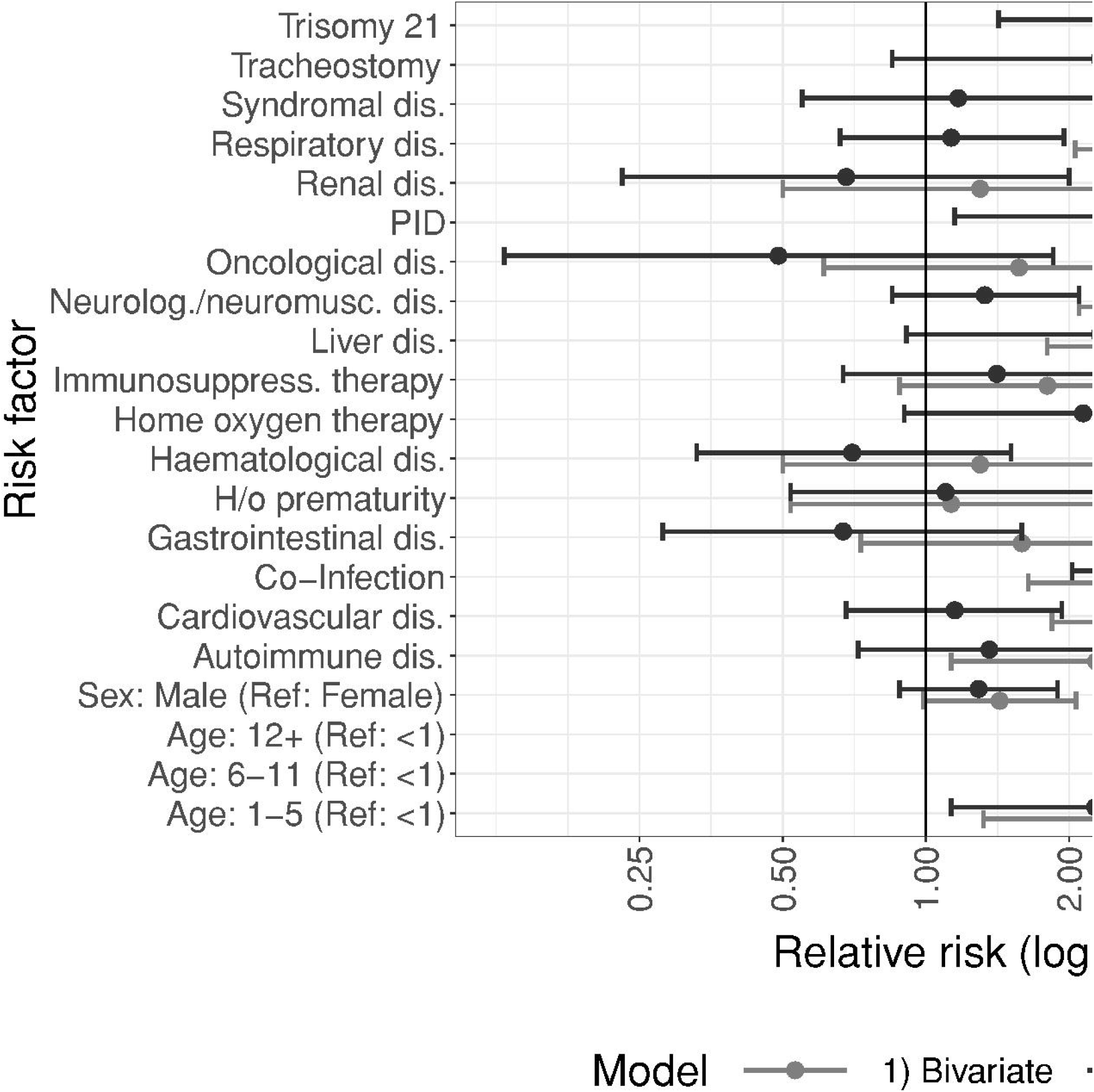
Predictors for ICU admission in bivariate and adjusted models. The adjusted model additionally accounts for ethnic background (Caucasian/Arabic/African/other). Coefficient estimates for ethnic background are not shown.

Due to the low number of sequelae and reported deaths, predictors of these outcomes could not be calculated.

## DISCUSSION

With nearly 50% of hospitals participating, as well as including data of a large proportion of children, hospitalized with SARS-CoV-2-infection, the DGPI registry has provided the means to reliably analyze the clinical spectrum of hospitalized children and adolescents with SARS-CoV-2 infections in Germany. The registry includes detailed information on clinical manifestations, demographic factors and predictors of disease severity. Therefore, our results are comprehensive and most likely to be applicable to countries with a similar medical and socioeconomic environment.

Our data confirm previous studies showing that SARS-CoV-2 infections in children usually are mild and that they are associated with a low rate of hospitalization and intensive care [11, 12]. Of note, more than 50% of registered children and adolescents were not admitted to hospital due to a SARS-CoV-2 infection, but rather for other reasons. In these cases, SARS-CoV-2 infection can be considered an accidental finding, even though the patients developed clinical signs of COVID-19 during their hospital stays. Furthermore, only 20% of patients received any SARS-CoV-2-related therapy during their hospital stays, suggesting that most hospitalization would have happened regardless of the SARS-CoV-2 infection. These observations emphasize that symptoms of SARS-CoV-2 infection are generally mild [13] for the majority of the pediatric population, even for those who become hospitalized.

As an interesting secondary finding, we discovered that although schools in Germany remained open during the first part of the country’s second pandemic wave (November through mid-December 2020). In half of the cases information on exposure was missing, however the vast majority of infections among children and adolescents were reported to be acquired within the household [14] — not in schools. This supports findings from others [15, 16] who have concluded that educational settings play only a minor role in SARS-CoV-2 transmission in comparison with other factors.

While low morbidity and mortality in the pediatric population in general is reassuring, our ability to identify relevant sub-populations who may have an increased risk of severe disease course remains critical — for the health outcomes of the patients themselves, as well as for the purpose of implementing targeted protection measures and in order to properly advise authorities on vaccine prioritization.

Although infants were hospitalized at a higher rate than older children relative to their share in the general population [12, 17], only 14% of infants required a SARS-CoV-2-related therapy. Interestingly, this is a significantly lower proportion than for older age groups. In addition, prematurely born infants were less likely to require therapy as compared to older children. However, this does not indicate that infants are more likely to experience a severe disease course. In fact, most admissions in this age group were likely due to precautionary measures in febrile neonates or young infants rather than to SARS-CoV-2 disease severity as such [4].

By using ICU admission as a surrogate parameter for disease severity, several comorbidities were significantly associated with the need for ICU admission in a bivariate model, only patient age, trisomy 21, coinfections and PIDs remained significant risk factors in the fully adjusted model. Remarkably, an underlying oncologic disease was not associated with increased risk of severe disease course. However, this may be explained by the fact that most children with ongoing cancer treatment are more meticulously mitigated and under closer surveillance for SARS-CoV-2 infection. Thus, more clinically asymptomatic infections may have been detected in this specific population, with patients more likely to be admitted to hospital for precautionary reasons.

The fact that the relative risk in our cohort was highest in the 6-to 11-year-old age group, and simultaneously decreased among adolescents, seems intriguing. It is, however, possible that this finding is biased by the fact that some adolescents could have been treated in adult care units. As a result, they may not have been included in this registry, because they were admitted to adult rather than children’s hospitals. Given the overall low number of hospitalized children and adolescents — as well as critically ill children— reliable prediction of outcome remains difficult.

Analyzing defined diseases, patients with pulmonary hypertension, cyanotic heart disease, s/p cardiac surgery, fatty liver disease, neuromuscular impairment and epilepsy seem to have a higher risk for severe disease. Consequently, children belonging to these groups, and/or who have these comorbidities, might also be at higher risk of severe disease and should therefore be taken into consideration concerning protection measures. This is particularly true for children and adolescents who are characterized by multiple risk factors.

Only patients with a SARS-CoV-2 infection laboratory-confirmed via PCR or rapid antigen test were included in this registry. Consequently, PIMS cases in Germany were included in this analysis only if the patient’s PCR and/or antigen test was still positive. This group, however, represents only a small fraction of all PIMS patients. Because the DGPI’s COVID registry was not specifically designed to identify PIMS cases, our group established a supplementary nationwide registry for this purpose [18]. Clearly, PIMS is an important complication of SARS-CoV-2 infection among children and needs to be considered as a significant SARS-CoV-2-associated morbidity. However, given that over 50% of patients are over six years of age and rarely have comorbidities, including PIMS in this analysis would have been unlikely to change our finding that older age within the pediatric age range is the strongest predictor for disease severity. Last, only a small proportion of children and adolescents with SARS-CoV-2 infection may go on to develop PIMS [19]. To date, no parameters are available to predict the development of PIMS following infection. Therefore, at least at this point in time, analysis of PIMS patients will not allow us to identify those who might benefit from infection-preventive measures, such as higher priority for vaccination.

### Limitations

The main limitation of our analysis is the potential selection bias of cases reported to the DGPI registry. Therefore, the relative risks for ICU admission refer to hospitalized children with SARS-CoV-2 detection only. Disease severity and relative risks might be different in children hospitalized due to COVID-19. Population-based estimations of disease severity based on statutory notification data and estimations of registry coverage are subject to underreporting and possibly reporting bias. Different age medians of hospitalized children within the statutory notification system and the DGPI registry might indicate an underrepresentation of adolescents in the registry potentially resulting from their treatment in adult care units. In addition, the overall low case numbers of pediatric patients treated in the ICU make reliable analysis challenging, especially when aiming to define single comorbidities. Further, end of patient follow-up with end of hospitalization may impair detection of long-term sequelae.

### Conclusion

Overall, a small proportion of children and adolescents was hospitalized in Germany during the first year of the pandemic. The majority of patients within our registry was not admitted due to COVID-19 suggesting an overestimation of the disease burden even in hospitalized children. Nevertheless, a large proportion of children and adolescents with confirmed COVID-19 reported in Germany could be captured. This allowed for detailed assessment of overall disease severity and underlying risk factors in our cohort. The main risk factors for COVID-19 disease associated intensive care treatment were older patient age, trisomy 21, PIDs and coinfection at the time of hospitalization. Pulmonary hypertension, cyanotic heart disease, s/p cardiac surgery, fatty liver disease, epilepsy and neuromuscular impairment also appear to increase risk for ICU admission. Hospitalization itself did not correspond to severe disease in our cohort given that 80% patients did not require SARS-CoV-2 related therapy.

### What is already known on this topic?

Children and adolescents usually have mild COVID-19 disease courses with low hospitalization rates.

Less information on risk factors and outcome predictors is available in children with SARS-CoV-2 infections compared to adults.

### Added value of this study?

Even in hospitalized children and adolescents 80% do not require SARS-CoV-2 related therapy.

Patient age in the pediatric population, trisomy 21, PIDs and coinfections are the main risk factors for a severe disease course.

Pulmonary hypertension, cyanotic heart disease, s/p cardiac surgery, fatty liver disease, epilepsy and neuromuscular impairment increase the risk for ICU admission, while asthma, diabetes and arterial hypertension do not.

This cohort has enabled identification of vulnerable populations who are likely to benefit from targeted protection measures and vaccine prioritization.

## Data Availability

We share data if reasonable requests are received. Requests should be directed to the corresponding author at

## Author Contributorship

J.A., M.H., J.H., A.S. and R.B. designed and started the registry; J.A., M.D. and N.D. managed the database and validated the data; DT.S., J.B. and A.T. coordinated resources; J.A., M.R. and J.S. analyzed the registry data; F.R., W.H., J.S. and S.H. collected and analyzed the national surveillance data; J.A., M.R., F.R. and M.D. wrote the original draft of the manuscript. M.H., N.D., W.H., J.S., S.H., J.H., A.S., DT.S. J.B., A.T., J.S. und R.B. reviewed and edited the manuscript.

## Ethical approval

The registry was approved by the Ethics Committee of the Technische Universität (TU) Dresden (BO-EK-110032020) and was assigned clinical trial number DRKS00021506.

## Acknowledgments

We thank each and every staff member in the participating hospitals for reporting their cases to the DGPI registry.

Furthermore, we would like to thank Doris Altmann for providing and Ann-Sophie Lehfeld for validating the presented statutory notification system data. We would like to extend our thanks to the local health authorities and the respective state health authorities, which collected this data, validated it locally and reported it to the Robert Koch Institute.

## Legend

### Supplemental Tables

**Supplemental Table 1**. Reported comorbidities in patients 0-17 y, 18 March 2020 – 30 April 2021 (statutory notification system).

## Abbreviations

CI: Confidence interval
COVID-19: Coronavirus disease 2019
RTA: Rapid antigen test
PID: Primary Immunodeficiency
ICU: Intensive care unit
h/o: History of
s/p: Status post
IQR: Interquartile Range
MIS-C: Multisystem Inflammatory Syndrome in Children
PCR: Polymerase Chain Reaction
PIMS: Pediatric Inflammatory Multisystem Syndrome
SARS-CoV-2: Severe acute respiratory syndrome Coronavirus type 2

## REFERENCES

1 Zhu N, Zhang D, Wang W, et al. A Novel Coronavirus from Patients with Pneumonia in China, 2019. N Engl J Med 2020;382(8):727–33. doi:10.1056/NEJMoa2001017 [published Online First: 24 January 2020].

2 Li J, Huang DQ, Zou B, et al. Epidemiology of COVID-19: A systematic review and meta-analysis of clinical characteristics, risk factors, and outcomes. J Med Virol 2021;93(3):1449–58. doi:10.1002/jmv.26424 [published Online First: 25 August 2020].

3 Mehta NS, Mytton OT, Mullins EWS, et al. SARS-CoV-2 (COVID-19): What Do We Know About Children? A Systematic Review. Clin Infect Dis 2020;71(9):2469–79.

4 Ouldali N, Yang DD, Madhi F, et al. Factors Associated With Severe SARS-CoV-2 Infection. Pediatrics 2021;147(3). doi:10.1542/peds.2020-023432 [published Online First: 15 December 2020].

5 Feldstein LR, Tenforde MW, Friedman KG, et al. Characteristics and Outcomes of US Children and Adolescents With Multisystem Inflammatory Syndrome in Children (MIS-C) Compared With Severe Acute COVID-19. JAMA 2021;325(11):1074–87.

6 Harris PA, Taylor R, Minor BL, et al. The REDCap consortium: Building an international community of software platform partners. J Biomed Inform 2019;95:103208. doi:10.1016/j.jbi.2019.103208 [published Online First: 9 May 2019].

7 Harris PA, Taylor R, Thielke R, et al. Research electronic data capture (REDCap)--a metadata-driven methodology and workflow process for providing translational research informatics support. J Biomed Inform 2009;42(2):377–81. doi:10.1016/j.jbi.2008.08.010 [published Online First: 30 September 2008].

8 RKI. Falldefinition Coronavirus-Krankheit-2019 (COVID-19) (SARS-CoV-2). https://www.rki.de/DE/Content/InfAZ/N/Neuartiges_Coronavirus/Falldefinition.pdf?blob=publicationFile (accessed 24 May 2021).

9 Statistisches Bundesamt Deutschland - GENESIS-Online: Tabelle abrufen NaN. Available at: https://www-genesis.destatis.de/genesis//online?operation=table&code=12411-0005&bypass=true&levelindex=0&levelid=1622145550488#abreadcrumb Accessed May 31, 2021.

10 Zou G. A modified poisson regression approach to prospective studies with binary data. Am J Epidemiol 2004;159(7):702–06.

11 Castagnoli R, Votto M, Licari A, et al. Severe Acute Respiratory Syndrome Coronavirus 2 (SARS-CoV-2) Infection in Children and Adolescents: A Systematic Review. JAMA Pediatr 2020;174(9):882–89.

12 Dong Y, Mo X, Hu Y, et al. Epidemiology of COVID-19 Among Children in China. Pediatrics 2020;145(6). doi:10.1542/peds.2020-0702 [published Online First: 16 March 2020].

13 He J, Guo Y, Mao R, et al. Proportion of asymptomatic coronavirus disease 2019: A systematic review and meta-analysis. J Med Virol 2021;93(2):820–30. doi:10.1002/jmv.26326 [published Online First: 13 August 2020].

14 Galow L, Haag L, Kahre E, et al. Lower household transmission rates of SARS-CoV-2 from children compared to adults. J Infect 2021. doi:10.1016/j.jinf.2021.04.022 [published Online First: 28 April 2021].

15 Hobbs CV, Martin LM, Kim SS, et al. Factors Associated with Positive SARS-CoV-2 Test Results in Outpatient Health Facilities and Emergency Departments Among Children and Adolescents Aged <18 Years - Mississippi, September-November 2020. MMWR Morb Mortal Wkly Rep 2020;69(50):1925–29. doi:10.15585/mmwr.mm6950e3 [published Online First: 18 December 2020].

16 Ismail SA, Saliba V, Lopez Bernal J, et al. SARS-CoV-2 infection and transmission in educational settings: a prospective, cross-sectional analysis of infection clusters and outbreaks in England. The Lancet Infectious Diseases 2021;21(3):344–53.

17 Swann OV, Holden KA, Turtle L, et al. Clinical characteristics of children and young people admitted to hospital with covid-19 in United Kingdom: prospective multicentre observational cohort study. BMJ 2020;370:m3249. doi:10.1136/bmj.m3249 [published Online First: 27 August 2020].

18 DGPI: Deutsche Gesellschaft für Pädiatrische Infektiologie. PIMS-Survey: Pediatric Inflammatory Multisystem Syndrome (PIMS) in Deutschland 2021. Available at: https://dgpi.de/pims-survey-anleitung/ accessed May 24, 2021.

19 Dufort EM, Koumans EH, Chow EJ, et al. Multisystem Inflammatory Syndrome in Children in New York State. N Engl J Med 2020;383(4):347–58. doi:10.1056/NEJMoa2021756 [published Online First: 29 June 2020].

