## supplemental table 1 for "Risk factors for hospitalization, disease severity and mortality in children and adolescents with COVID-19: Results from a nationwide German registry"

### Supplemental Table 1.
### Reported comorbidities in patients 0-17 y, 18 March 2020 – 30 April 2021 (statutory notification system)

| | **Non-Hospitalized N= 222117** | | **Hospitalized N= 3354** | |
|---|---|---|---|---|
| | *Yes (n)* | *%* | *Yes (n)* | *%* |
| Respiratory disease | 4305 | 1.9 | 108 | 3.2 |
| Cardiovascular disease | 1405 | 0.6 | 99 | 3.0 |
| Renal disease | 439 | 0.2 | 36 | 1.1 |
| Liver disease | 119 | 0.1 | 11 | 0.3 |
| Oncologic disease | 232 | 0.1 | 44 | 1.3 |
| Autoimmune disease | 402 | 0.2 | 41 | 1.2 |

*Frequencies and percentages based on number of cases for which hospitalization status and information on comorbidity was reported.*
